# Mendelian Randomization Analysis of the Relationship Between Native American Ancestry and Gallbladder Cancer Risk

**DOI:** 10.1101/2022.05.03.22274595

**Authors:** Linda Zollner, Felix Boekstegers, Carol Barahona Ponce, Dominique Scherer, Katherine Marcelain, Valentina Gárate-Calderón, Melanie Waldenberger, Erik Morales, Armando Rojas, César Munoz, Bettina Müller, Javier Retamales, Gonzalo de Toro, Allan Vera Kortmann, Olga Barajas, María Teresa Rivera, Analía Cortés, Denisse Loader, Javiera Saavedra, Lorena Gutiérrez, Alejandro Ortega, Maria Enriqueta Bertrán, Leonardo Bartolotti, Fernando Gabler, Mónica Campos, Juan Alvarado, Fabricio Moisán, Loreto Spencer, Bruno Nervi, Daniel Carvajal, Héctor Losada, Mauricio Almau, Plinio Fernández, Jordi Olloquequi, Alice R. Carter, Juan Francisco Miquel Poblete, Bernabe Ignacio Bustos, Macarena Fuentes Guajardo, Rolando Gonzalez-Jose, Maria Cátira Bortolini, Victor Acuña-Alonzo, Carla Gallo, Andres Ruiz Linares, Francisco Rothhammer, Justo Lorenzo Bermejo

## Abstract

**Background:** A strong association between the proportion of Native American ancestry and the risk of gallbladder cancer (GBC) has been reported in observational studies. Chileans show the highest incidence of GBC worldwide, and the Mapuche are the largest Native American people in Chile. We set out to investigate the causal association between Native American Mapuche ancestry and GBC risk, and the possible mediating effects of gallstone disease and body mass index (BMI) on this association.

**Methods:** Markers of Mapuche ancestry were selected based on the informativeness for assignment measure and then used as instrumental variables in two-sample mendelian randomization (MR) analyses and complementary sensitivity analyses.

**Result:** We found evidence of a causal effect of Mapuche ancestry on GBC risk (inverse variance-weighted (IVW) risk increase of 0.8% for every 1% increase in Mapuche ancestry proportion, 95% CI 0.4% to 1.2%, p = 6.6×10^-5^). Mapuche ancestry was also causally linked to gallstone disease (IVW risk increase of 3.6% per 1% increase in Mapuche proportion, 95% CI 3.1% to 4.0%, p = 1.0×10^-59^), suggesting a mediating effect of gallstones in the relationship between Mapuche ancestry and GBC. In contrast, the proportion of Mapuche ancestry showed a negative causal effect on BMI (IVW estimate -0.006 kg/m^2^ per 1% increase in Mapuche proportion, 95% CI -0.009 to -0.003, p = 4.4×10^-5^).

**Conclusions:** The results presented here may have significant implications for GBC prevention and are important for future admixture mapping studies. Given that the association between Mapuche ancestry and GBC risk previously noted in observational studies appears to be causal, primary and secondary prevention strategies that take into account the individual proportion of Mapuche ancestry could be particularly efficient.

## Introduction

Every year, around 116,000 people are diagnosed with gallbladder cancer (GBC) and 85,000 die due to this aggressive disease worldwide (The Global Cancer Observatory, 2020). The malignancy of the biliary tract primarily affects women in low- and middle-income countries, and relatively little effort has been invested in research on GBC (Mehrotra et al., 2018; Stinton & Shaffer, 2012).

The development of GBC is probably driven by a combination of environmental exposures and genetic predisposition (Hundal & Shaffer, 2014). Symptoms are often absent or unspecific until the disease has progressed to a non-curative stage, leaving patients with few treatment options (Hundal & Shaffer, 2014; Roa & de Aretxabala, 2015; Stinton & Shaffer, 2012). As GBC is mostly diagnosed at an advanced stage, the 5-year survival rate is low, reported at between 5% and 30% depending on the country of origin of the study population (Bertran et al., 2010; Hundal & Shaffer, 2014; Kanthan et al., 2015; Stinton & Shaffer, 2012; Zhu et al., 2020). Gallbladder removal (cholecystectomy) is a valuable tool for primary and secondary prevention of GBC, but little progress has been made in individualized risk prediction and early diagnosis (Roa & de Aretxabala, 2015).

The incidence of GBC shows wide geographical and ethnic variation (Mehrotra et al., 2018). High-income regions such as Western Europe, the United States and Australia have 1‒2 cases per 100,000 person-years. In contrast, the largest Native American people in Chile – the Mapuche – shows the highest incidence of GBC in the world, with more than 20 cases per 100,000 person-years (Campbell et al., 2017). Observational studies have found a strong association between the individual proportion of Native American Mapuche ancestry and GBC risk: each 1% increase in the proportion of Mapuche ancestry translates into a 3.7% increase in GBC mortality (Lorenzo Bermejo et al., 2017). However, the observed association may arise from other established GBC risk factors, especially gallstones and elevated body mass index (BMI) (Hundal & Shaffer, 2014; Roa & de Aretxabala, 2015; Stinton & Shaffer, 2012). The Mapuche have a high prevalence of gallstone disease, a relative GBC risk of 4.9 has been observed in individuals with a history of gallstones, and recent studies have found evidence of a causal effect of gallstones on GBC risk for genetically admixed Chileans (odds ratio [OR] = 1.97) (Barahona Ponce et al., 2021; Mehrotra et al., 2018; Stinton & Shaffer, 2012). Native American ancestry has also been associated with increased BMI: the World Cancer Research Fund considers that higher body fatness marked by BMI probably causes GBC, and relative GBC risks of 1.59 for women and 1.09 for men per five-point increase in BMI have been reported (Campbell et al., 2017; Hundal & Shaffer, 2014; Roa & de Aretxabala, 2015; World Cancer Research Fund/American Institute for Cancer Research, 2018). Furthermore, multiparity in women, low socio-economic status, chronic inflammation and lifestyle choices such as cigarette smoking and alcohol consumption could also confound the association between ethnicity and GBC, justifying the use of MR to assess the causal relationship between Mapuche ancestry and GBC risk (Lugo et al., 2020; Roa & de Aretxabala, 2015; Stinton & Shaffer, 2012; Wistuba & Gazdar, 2004).

In response to the high GBC mortality rates and lack of treatment options, but based on weak scientific evidence, the Chilean government is implementing a GBC prevention program that provides financial support to gallstone patients for prophylactic cholecystectomy (Roa & de Aretxabala, 2015). Prioritizing prophylactic cholecystectomy for those at high risk of GBC, taking into account causal risk factors rather than observational associations potentially attributable to confounding, would optimize the efficiency of current GBC prevention measures. Mapuche ancestry per se neither causes nor prevents disease, but statistical inference on the causal effect of the individual’s proportion of Mapuche ancestry on GBC risk allows for a distinction between correlation and confounding-free causation, allowing for prioritization of causal risk factors for prevention and a better understanding of the underlying etiological mechanisms leading to GBC development.

With this goal in mind, we preselected markers of Mapuche ancestry in a panel of Native American, European and African individuals, relying on the informativeness for assignment measure (Rosenberg et al., 2003). We then used a subset of ancestry markers as instrumental variables for the individual proportion of Mapuche ancestry in two-sample MR to assess the causal effect of Mapuche ancestry on GBC risk. It should be noted that population stratification, typically a limitation in MR studies, was the exposure of interest in the present study.

## Results

The flowchart in **Figure 1** describes the performed analyses with some intermediate results. Preselection of markers of Mapuche ancestry in a reference panel composed of Native American Mapuche (n = 28), Native American Aymara (n = 63), European (CEU and IBS, n = 206) and African (YRI, n = 108) individuals relying on the informativeness for assignment measure yielded 21,854 candidate markers. Of these, 985 genetic variants were robustly associated (p < 5×10^-8^) with the individual proportion of Mapuche ancestry, which was previously estimated in a dataset with genome-wide genotype data from 1,861 genetically admixed Chileans. Exclusion of markers associated with established GBC risk factors (PheWAS p < 5×10^-8^) complemented with LD pruning (r^2^ < 0.01) resulted in 430 instrumental variables preliminarily retained for the subsequent MR analyses. Radial MR based on summary statistics of the association between the instrumental variables and the proportion of Mapuche ancestry (sample I: 1,861 admixed Chileans) and that between the instrumental variables and GBC status (sample II: 412 Chilean GBC patient‒control pairs), both summary statistics adjusted for age and sex, detected 33 outlying instruments. The subsequent MR analysis of the association Mapuche ancestry → GBC was therefore based on 397 instrumental variables, which explained 13.2% of the variance in the proportion of Mapuche ancestry.

**Figure 1.**
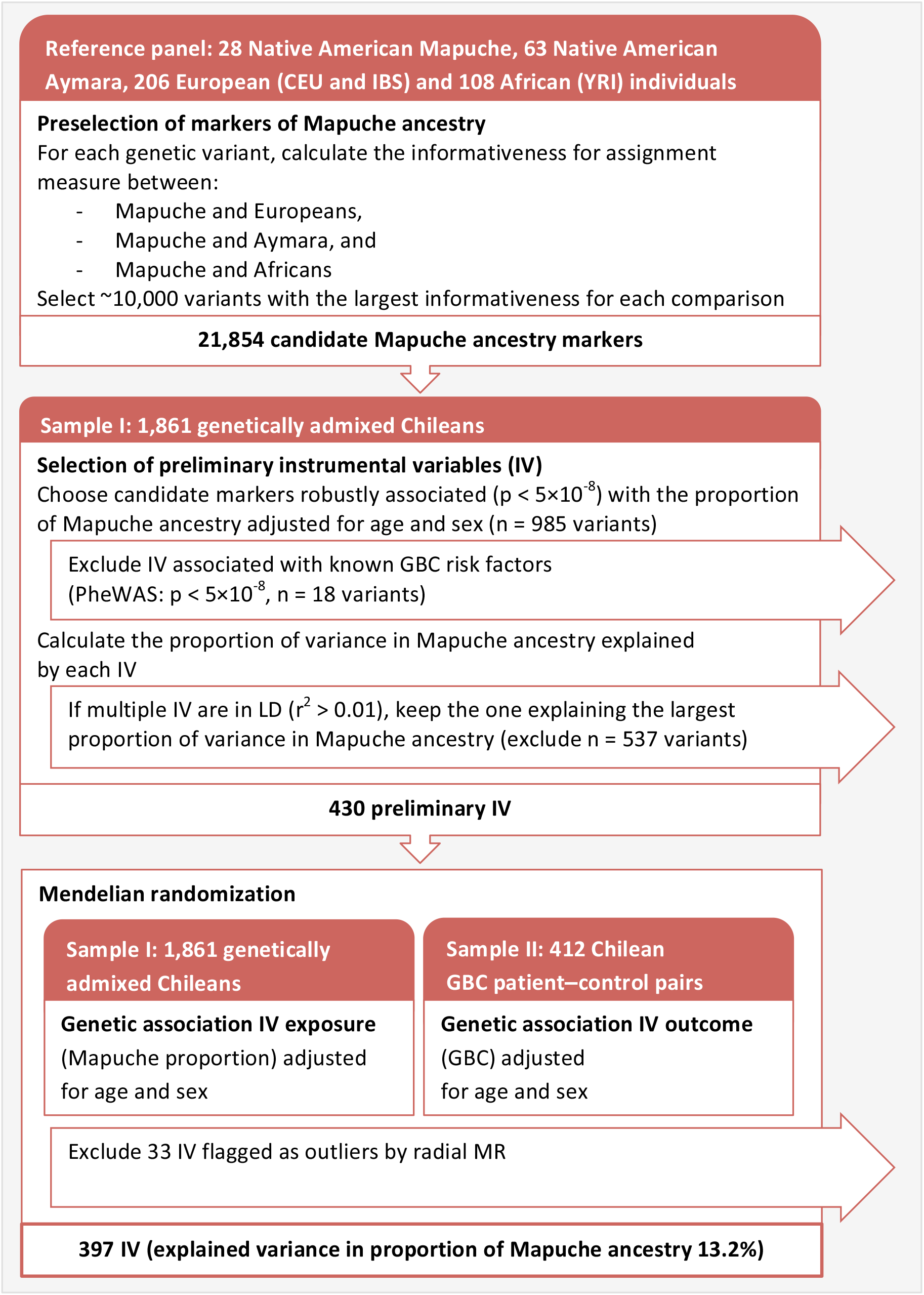
Flowchart describing the main analyses, from the selection of markers of Mapuche ancestry in a reference panel composed of Mapuche, Aymara, European and African individuals, to the two-sample MR based on 397 selected instrumental variables after exclusion of outlying instruments based on radial MR

**Figure 2** depicts the genetic principal components, and the estimated proportions of Mapuche, Aymara and European ancestry in the two samples used for the main MR analysis. Individuals with a proportion of Mapuche ancestry above the 95^th^ percentile in the sample are represented in orange; blue and green highlight individuals with the largest proportions of Aymara and European ancestry, respectively. The first principal component in admixed Chileans distinguished between European and Native American ancestry, and the second principal component separated the two main types of Native American ancestry in Chile: Mapuche and Aymara. A strong correlation between the second principal component and the proportion of Mapuche ancestry was found in the two samples used for MR analysis (panels C and F in **Figure 2**). In the three plots depicting sample II (panels D, E and F in **Figure 2**), crosses represent GBC patients and circles represent population-based controls. The concentration of GBC patients among individuals with the highest proportion of Native American ancestry, especially Mapuche ancestry, was striking.

**Figure 2.**
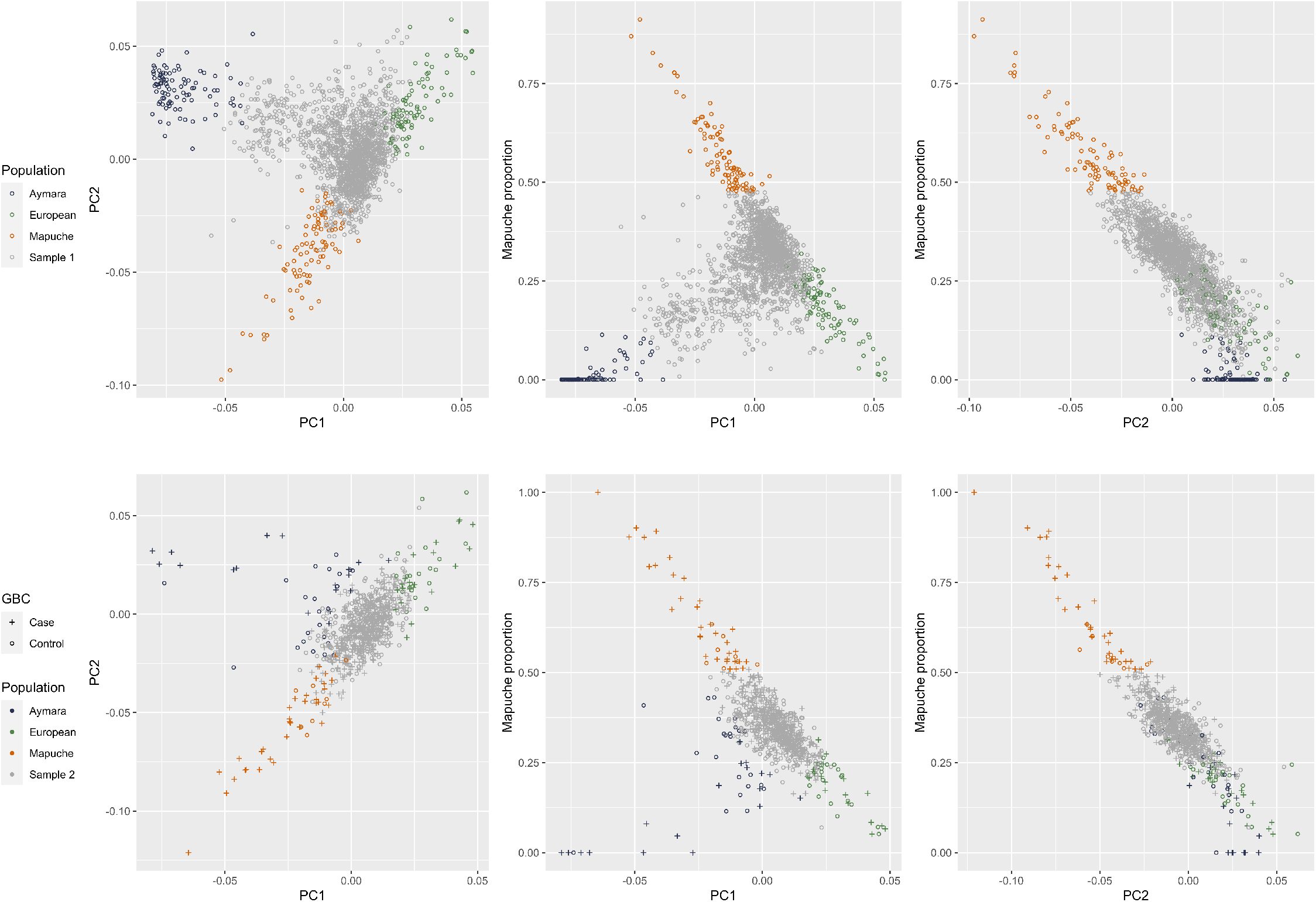
Scatter plots of first and second genetic principal components (PC), and the individual proportion of Mapuche ancestry. Individuals with proportions of Mapuche, Aymara and European ancestry above the 95th percentile are shown in orange, blue and green, respectively. Panels A, B and C refer to sample I; panels D, E and F refer to sample II used in the main MR analysis, respectively. In sample II, GBC patients are represented by crosses and population-based controls by circles.

MR analysis of the association Mapuche ancestry → GBC risk revealed no heterogeneity among instrumental variables as a proxy for pleiotropy (inverse variance-weighted (IVW) Cochran’s *Q* statistic p = 0.99) and no horizontal pleiotropy (MR-Egger intercept p = 0.87; **Table 1**). Neither outliers nor weak instrument biases were apparent in the scatter and funnel plots (panels A and B in **Figure 3**). We found evidence of a causal effect of Mapuche ancestry on GBC risk (IVW OR = 1.008, which translates into a GBC risk increase of 0.8% for every 1% increase in the proportion of Mapuche ancestry, 95% confidence interval [CI] 0.4% to 1.2%, p = 6.6×10^-5^). MR-Egger (OR = 1.009, 95% CI 1.004 to 1.015, p = 6.7×10^-4^) and weighted median estimates (OR = 1.009, 95% CI 0.991 to 1.028, p = 0.33) were consistent with IVW estimates. As described above, the second principal component of genetic variability in admixed Chileans reflects the individual proportion of Mapuche ancestry, and, as expected, the causal effect vanished after including the second principal component in the calculation of summary statistics (p = 0.62). In contrast, consideration of the first and third to tenth principal components translated into stronger estimates of the causal effect (1.4% risk increase, 95% CI 0.8% to 1.9%, p = 5.9×10^-7^).

**Table 1.**
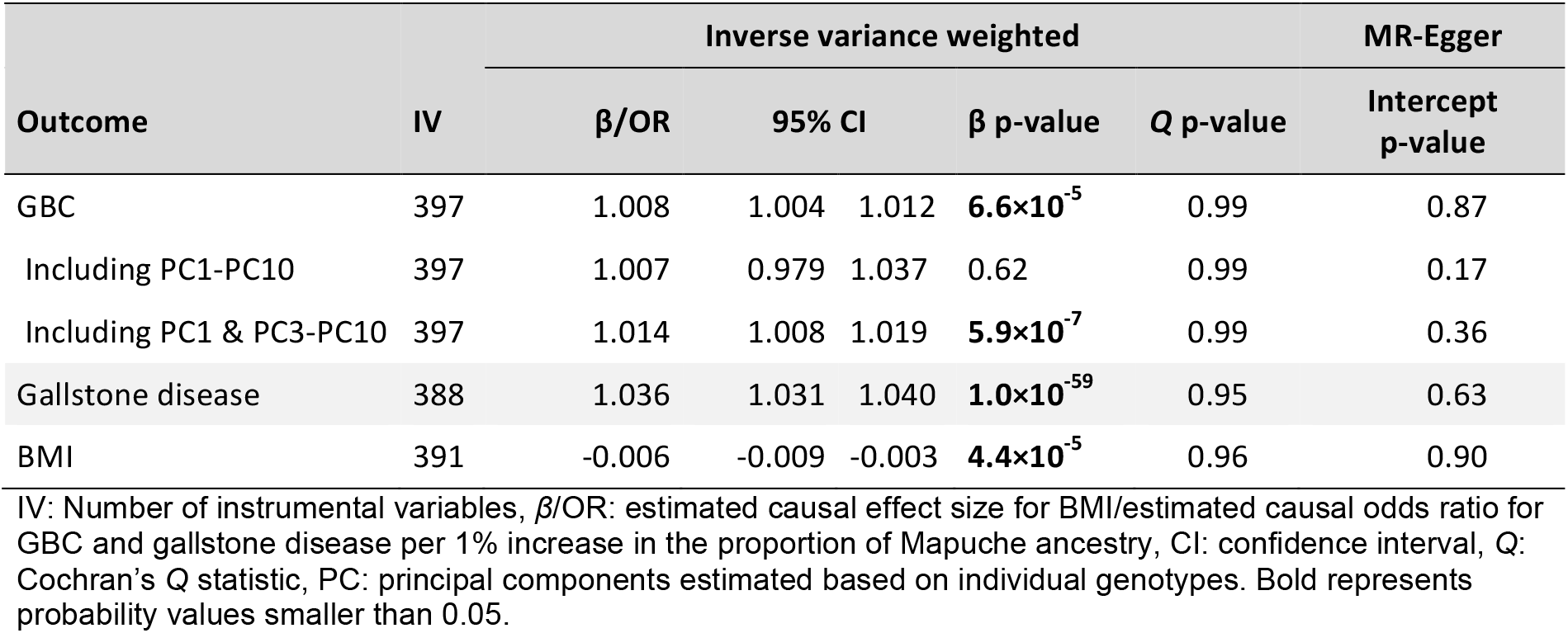
MR results on the causal relationship between Native American Mapuche ancestry as exposure and GBC, gallstone disease and BMI as outcomes of interest. Cochran’s *Q* statistic p-values higher than 0.05 are suggestive of no instrument heterogeneity as a proxy for pleiotropy, and MR-Egger intercept p-values higher than 0.05 are consistent with no horizontal pleiotropy. Results from sensitivity analyses on the association Mapuche ancestry → GBC risk after including the first to the tenth principal component, both with and without the second principal component, which reflects the individual proportion of Mapuche ancestry in admixed Chileans, are also shown.

**Figure 3.**
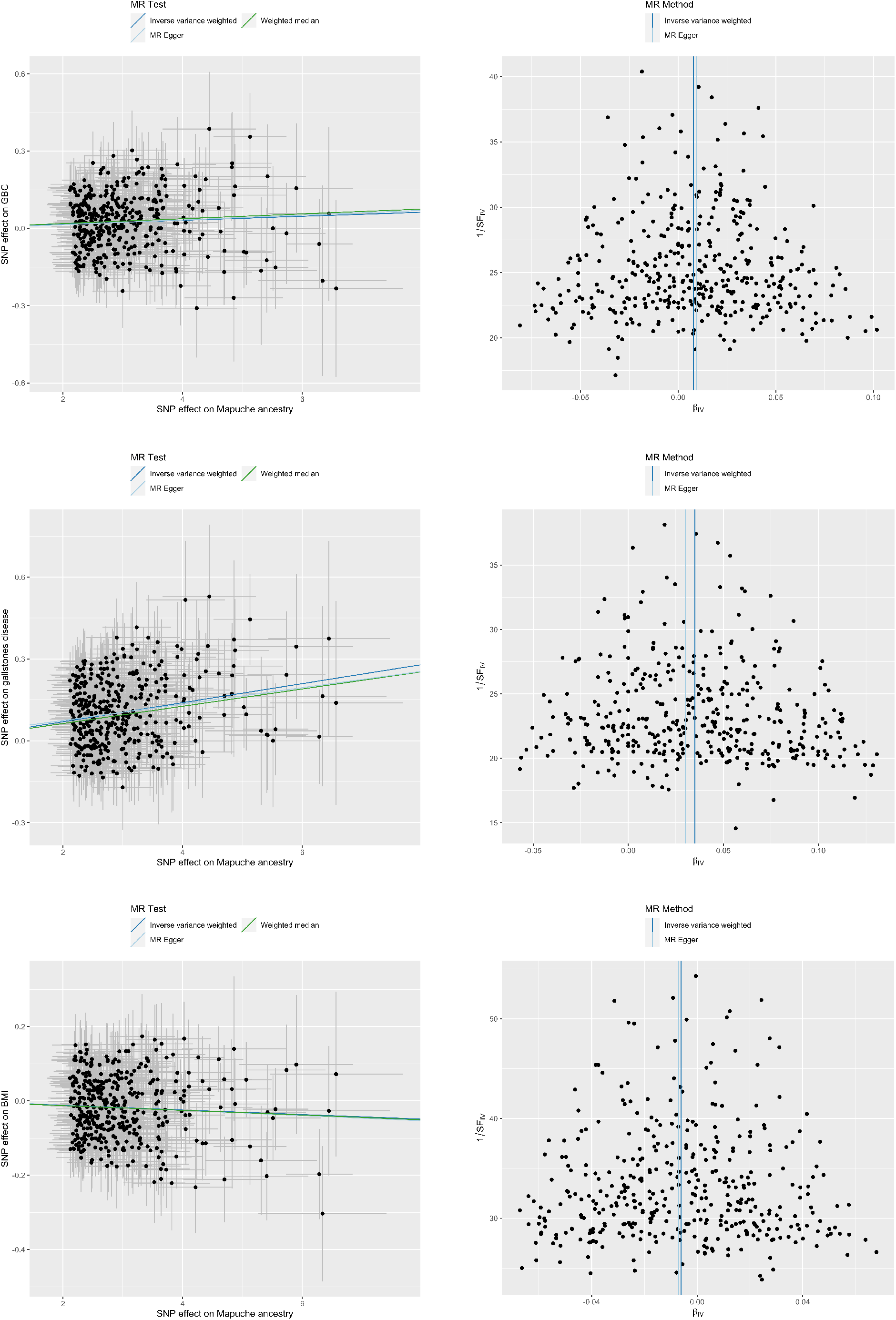
Scatter and funnel plots from MR analyses after exclusion of outlying instruments according to radial MR on the association between Mapuche ancestry and GBC risk (panels A and B), gallstone disease (panels C and D) and BMI (panels E and F)

Further sensitivity analysis considered two-step MR and multivariable MR (MVMR) to investigate the potential mediating effects of gallstone disease and BMI on the link between Mapuche ancestry and GBC. For two-step MR mediation analysis, we first assessed the causal effect of Mapuche ancestry on gallstone disease. Neither heterogeneity among instrumental variables (*Q* statistic p = 0.95) nor horizontal pleiotropy (MR-Egger intercept p = 0.63) was noted. We found evidence of a causal effect of Mapuche ancestry on the risk of gallstone disease (IVW risk increase of 3.6% for every 1% increase in Mapuche proportion, p = 1.0×10^-59^; MR-Egger OR = 1.032, 95% CI 1.026 to 1.039, p = 5.2×10^-24^; weighted median OR = 1.031, 95% CI 1.010 to 1.051, p = 0.003). These results are visually represented in **Figure 3** (panels C and D).

Since we found a causal effect of Mapuche ancestry on gallstones in the present study, and a causal effect of gallstones on GBC risk has already been demonstrated for Chileans, the two-step MR analysis suggests that gallstones mediate the association between Mapuche ancestry and GBC risk. To validate this hypothesis, we also performed MVMR. The instrumental variables used for MVMR were weak (Mapuche ancestry F statistics = 3.49; gallstones F statistics = 1.37), but horizontal pleiotropy was not found (*Q* statistic of 425 on 432 degrees of freedom, p = 0.59). In agreement with the results of two-step MR, MVMR revealed a causal effect of gallstones on GBC risk (OR = 1.263, p = 2.0×10^-9^) but no concurrent causal effect of Mapuche ancestry on GBC risk (OR=1.0001, p = 0.95).

Neither heterogeneity among instrumental variables (*Q* statistic p = 0.96) nor horizontal pleiotropy (MR-Egger intercept p = 0.90) was detected in MR analysis of the association Mapuche ancestry → BMI. We found evidence of a negative causal effect of Mapuche ancestry on BMI (IVW *β* = -0.006 kg/m^2^ per 1% increase in Mapuche proportion, 95% CI -0.009 to -0.003, p = 4.4×10^-5^). The corresponding scatter and funnel plots are shown in panels E and F of **Figure 3**. MR-Egger (*β* = -0.007 kg/m^2^ per 1% Mapuche ancestry proportion, 95% CI -0.020 to 0.006, p = 0.31) and weighted median estimates (*β* = -0.006 kg/m^2^ per 1% Mapuche ancestry proportion, 95% CI -0.011 to -0.002, p = 3.8×10^-3^) showed good agreement with the negative causal effect estimated by IVW.

## Discussion

Utilizing ancestry informative markers as instrumental variables for the individual proportion of Mapuche ancestry in admixed Chileans, we applied two-sample MR to assess the causal relationship between Mapuche ancestry and GBC risk as the primary outcome, as well as two-step and MVMR to examine the mediating effects of gallstones and BMI on the relationship Mapuche ancestry → GBC risk. According to the MR results, the proportion of Mapuche ancestry is causally associated with GBC development and gallstones mediate this causal association. The practical implication of the identified causal relationships is that intensified surveillance and prophylactic gallbladder removal in gallstone carriers with a high percentage of Mapuche ancestry may improve the efficiency of current GBC prevention programs. The extent to which gallstones mediate the underlying mechanisms of the relationship between Mapuche ancestry and GBC needs to be quantified in future larger studies. Given the causal association between increasing proportions of Mapuche ancestry and decreasing BMI, and the causal association between increasing BMI and increasing GBC risk, it seems rather unlikely that BMI positively mediates between Mapuche ancestry and GBC risk.

Gallstones are highly prevalent in Chileans and one of the most important risk factors for GBC: observational studies report a relative GBC risk of 9.2‒10.1 for individuals with gallstones larger than 3 cm (Miquel et al., 1998; Stinton & Shaffer, 2012). We therefore examined the possible mediating effect of gallstone disease on the association between Mapuche ancestry and GBC risk. The evidence for a causal effect of Mapuche ancestry on gallstone disease was quite robust: the estimated causal OR was 1.036 per 1% Mapuche ancestry proportion. This finding, together with the previously reported causal effect of gallstones on GBC risk in Chileans, support a mediating effect of gallstones based on two-step MR (Barahona Ponce et al., 2021). The MVMR results, based on a relatively small sample size and weak instrumental variables, showed good agreement with two-step MR and also supported a mediating effect of gallstones between Mapuche ancestry and GBC risk; however, large prospective Chilean datasets with complete information on GBC, gallstones and individual genotype data are urgently needed to conduct formal mediation analyses and accurately quantify the magnitude of the direct and indirect effects of gallstones, which are highly relevant for more precise GBC prevention.

The International Agency for Research on Cancer, the American Institute for Cancer Research and the World Cancer Research Fund all consider obesity to be a likely cause of GBC, and high BMI is another important GBC risk factor, with a direct effect on GBC risk reported for Chileans and an indirect effect mediated by gallstones for Europeans (Barahona Ponce et al., 2021; Lauby-Secretan et al., 2016; World Cancer Research Fund/American Institute for Cancer Research, 2018). However, studies of the association between Native American ancestry and BMI have yielded contradictory results. BMI decreased by 0.13 m/kg^2^ for each 10% increase in the proportion of Native American ancestry in Mexican‒American women (Hu et al., 2015), and the association was also negative in a Hispanic‒Mexican study (Tang et al., 2006). In contrast, BMI increased by 0.56 m/kg^2^ per 10% increase in the proportion of Native American ancestry in Native Americans (Norden-Krichmar et al., 2014). Native American ancestry was positively correlated with obesity in Mexico and Peru, whereas no association was found in Brazil, Chile, and Colombia, and higher Native American ancestry was associated with overweight and obesity, but only among foreign-born Latina women (Ziv et al., 2006). In this study we found clear evidence of a negative causal effect of Mapuche ancestry on BMI, probably ruling out a positive mediating effect of BMI on the association between Mapuche ancestry and GBC risk.

The relatively small number of GBC patients investigated was a limitation of this study, especially considering the large sample sizes normally required for MR. However, the variance in the proportion of Mapuche ancestry explained by the instrumental variables was high (13.2%), and observational studies have reported a strong association between Mapuche ancestry and GBC risk. Besides low statistical power, another common limitation of MR studies is pleiotropy. First-order inverse variance weights keep the type I error rate under the causal null, and we calculated Cochran’s *Q* statistic using first-order weights to detect heterogeneity, which often reflects pleiotropy. We also visually inspected scatter and funnel plots, performed MR-Egger regression to detect potential bias attributable to horizontal pleiotropy, used radial MR to detect outlying instruments, and excluded instrumental variables associated with GBC risk factors not investigated in our study.

Previous studies have demonstrated the importance of subdividing Native American ancestry into its main subcomponents –in the case of admixed Chileans, the two major Native American subcomponents are Mapuche and Aymara ancestry (Lorenzo Bermejo et al., 2017). While combined Native American ancestry showed no association with GBC mortality and the proportion of Aymara ancestry showed a negative association with GBC mortality, each 1% increase in the Mapuche proportion translated into a 3.7% increase in the risk of death due to GBC (95% CI 3.1 to 4.3%). To assess the contribution of health system access to the identified association between Mapuche ancestry and GBC risk, hospitalization rates due to gallbladder removal (cholecystectomy) were included as an additional explanatory variable in the fitted regression models; this showed a minor role of health system access in the estimated increase in GBC mortality risk. The MR-based OR estimated in the present study confirmed a causal effect of Mapuche ancestry on GBC risk, while reducing the effect size to 0.8% (95% CI 0.4% to 1.2%). This result suggests that GBC prevention strategies that consider the individual proportion of Mapuche ancestry may be particularly efficient, and that the difference between the observational (3.7%) and causal (0.8%) increase in GBC risk is due to confounders of the association between Mapuche ancestry and GBC risk, which may include modifiable risk factors. Further research is warranted.

Beyond the practical relevance for GBC prevention, the results presented here may also be useful for designing future studies. On the one hand, the evidence of a causal effect of Mapuche ancestry on GBC risk underpins the potential of admixture mapping to identify novel GBC susceptibility variants, possibly in combination with subsequent association testing (note that the genome-wide significance level is much higher for admixture mapping than for association mapping) (Horimoto et al., 2021). For example, assuming that the average proportion of Mapuche ancestry in Chileans is about 40%, approximately 1100 GBC patients are needed to detect a Mapuche haplotype with a risk ratio of 1.8, consistent with the estimated causal OR of 1.008 per 1% proportion of Mapuche ancestry in the whole Chilean genome (McKeigue, 2005). On the other hand, the 3.7% increased GBC mortality risk per 1% Mapuche ancestry proportion previously reported in observational studies probably overestimates the contribution of Mapuche ancestry to GBC risk and would lead to underpowered admixture mapping studies. From an implementation perspective, recruiting study participants from the southern regions of Chile would increase the average proportion of Mapuche ancestry and thus the statistical power of the study.

In summary, we assessed the relationship between the individual proportion of Native American Mapuche ancestry and GBC risk using mendelian randomization. To our knowledge, this is the first MR study that considers genetic ancestry as the exposure of interest – it is important to keep in mind that the study was based on genetically admixed Chileans, who show continuous gradients of ancestry. Since statistical evidence of a causal effect of ancestry on disease development is generally more relevant than an observational association, which could also be due to confounding, the present findings provide refined information on the potential of accounting for ethnic differences (in this case Mapuche ancestry) in prevention of disease (in this case gallbladder cancer). While ancestry proportions are not modifiable exposures, personalized preventive measures could prioritize gallstone screening and prophylactic cholecystectomy for individuals with a high percentage of Mapuche ancestry. Future admixture mapping studies could also benefit from the methodology applied in the present investigation to test and quantify the causal effects of genetic ancestry on disease outcomes. From a more applied point of view, we found strong evidence for a causal effect of Mapuche ancestry proportion on GBC risk, most likely mediated by gallstones, with direct implications for the development of more efficient GBC prevention strategies.

## Materials and Methods

### Ethics approval

The study protocol conformed to the ethical guidelines of the 1975 Declaration of Helsinki and was approved by the ethics committees of Servicio de Salud Metropolitano Oriente, Santiago de Chile (#06.10.2015, #08.03.2016 and #12.11.2019), Servicio de Salud Metropolitano Sur Oriente, Santiago de Chile (#15.10.2015 and #05.04.2018), Servicio de Salud Metropolitano Central, Santiago de Chile (#1188-2015), Servicio de Salud Coquimbo, Coquimbo, Chile (#01.04.2016), Servicio de Salud Maule, Talca, Chile (#05.11.2015), Universidad Católica del Maule, Talca, Chile (#102-2020), Servicio de Salud Concepción, Concepción, Chile (ID: 16-11-97 and ID:19-12-111), Servicio de Salud Araucanía Sur, Temuco, Chile (#10.02.2020), Servicio de Salud Valdivia, Valdivia, Chile (ID:438), Centro de Bioética, Universidad del Desarrollo, Clínica Alemana de Santiago, Santiago de Chile (#2018-97, ID 678) and Unidad de Investigación Hospital San Juan de Dios, Santiago de Chile (#6182), the Medical Faculties of Universidad de Chile (approval #123-2012 and #11.10.2012) and Pontificia Universidad Católica de Chile (#11-159), and Universidad de Tarapacá and University College London as described in (Ruiz-Linares et al., 2014). All participants provided written informed consent before enrolment.

### Study participants

The Chilean genome is a mixture of chromosomal segments from two major Native American peoples, the Aymara in the north and the Mapuche in the south of the country; Europeans; and, to a lesser extent, Africans (Eyheramendy et al., 2015; Hundal & Shaffer, 2014; Ruiz-Linares et al., 2014). The reference panel for preselection of ancestry-informative markers and estimation of individual ancestry proportions in genetically admixed Chileans therefore included 63 Aymara (Reich et al., 2012) and 28 Mapuche (Lindo et al., 2018; Reich et al., 2012), as well as 206 Europeans (99 Utah residents with Northern and Western European ancestry [CEU] and 107 Iberians from Spain [IBS]), and 108 African Yorubans from Ibadan, Nigeria (YRI) from the 1000 Genome Project (The 1000 Genomes Project Consortium, 2015).

Sample I for two-sample MR analyses of the associations Mapuche ancestry → GBC, Mapuche ancestry → gallstone disease and Mapuche ancestry → BMI consisted of 1,861 Chileans from the Consortium for the Analysis of the Diversity and Evolution of Latin America (CANDELA) (Ruiz-Linares et al., 2014). Sample II for MR analysis of the association Mapuche ancestry → GBC risk was composed of 412 Chilean patients diagnosed with GBC and 412 population-based controls matched by age and sex with the GBC patients. Sample II for MR analysis of the association Mapuche ancestry → gallstone disease was composed of 351 Chilean gallstone patients and 351 controls matched by age and sex. Among GBC patients, 77% were diagnosed after surgical removal of the gallbladder (cholecystectomy), and gallstones were found in about 86% of the GBC patients investigated. Gallstone patients were patients who underwent cholecystectomy due to symptomatic gallstones. Population-based controls were selected from the Chilean subgroup of CANDELA and from Chilean studies on COPD and Chagas disease, with GBC and gallstone incidences representative of the general Chilean population (Blandino et al., 2022). Sample I and sample II for MR analyses on Mapuche ancestry → GBC risk and Mapuche ancestry → gallstone disease partially overlapped: 84 of the 412 controls and 91 of the 351 controls in the respective sample II also belonged to sample I. Sample II for MR analysis of the association Mapuche ancestry → BMI was based on 12,216 individuals from the Hispanic Community Health Study/Study of Latinos (HCHS/SOL, dbGaP accession number phs000810.v1.p1).

### Preselection of markers of Mapuche ancestry

To preselect genetic variants robustly associated with our exposure of interest, the individual proportion of Mapuche ancestry, we first chose markers of Mapuche ancestry based on the informativeness for assignment measure *I*_*n*_ (Rosenberg et al., 2003). For each genetic variant with *j* = *1, …, N* possible alleles in *i* = *1, …, K* subpopulations we calculated:

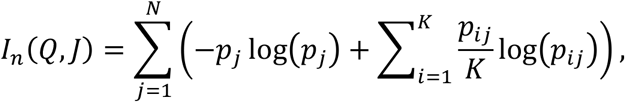

where *p*_*j*_ denotes the average frequency of allele *j* in all *K* subpopulations, *p*_*ij*_ the average frequency of allele *j* in subpopulation *i, Q* the (random) assignment of an individual to a subpopulation and *J* the (random) genotype of one of the two alleles of an individual. *I*_*n*_ conditioning on *Q* and *J* is a general approach to quantify the amount of information that multi-allelic markers provide about individual ancestry composition. We used the reference panel described above to estimate *I*_*n*_ for each of the 43,625 genetic variants available for all individuals in this study considering two subpopulations at a time (Mapuche – European, Mapuche – Aymara and Mapuche – African). We selected the 10,000 genetic variants with the highest *I*_*n*_ for each comparison and retained the genetic variants that were present in at least one comparison for the subsequent analyses.

### Selection of instrumental variables

In order to ensure that preselected ancestry informative markers fulfilled the first MR assumption (relevance), we used the software ADMIXTURE (version 1.3) for supervised estimation of the individual proportions of Mapuche ancestry in sample I, relying on the reference panel of Aymara, Mapuche, European and African individuals (Alexander et al., 2009), and retained as instrumental variables (IV) the genetic variants that showed a robust association (p < 5×10^-8^) with the estimated proportion of Mapuche ancestry, adjusted for age and sex. The second MR assumption (independence) was assessed by a phenome-wide association study (PheWAS), excluding genetic variants associated (p < 5×10^-8^) with potential confounders (menopause, educational level, diabetes, body circumference, smoking, alcohol consumption or gallstones) in MR-Base (Hemani et al., 2018). We then used the summary statistics of the association between the genetic variants and the estimated proportion of Mapuche ancestry (*β*) and the minor allele frequency (*MAF*) to calculate the variance in Mapuche ancestry proportion explained by each variant:

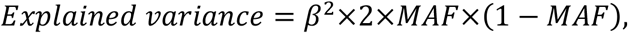

and excluded variants in linkage disequilibrium (LD) (r^2^ > 0.01) with other variants that explained a larger variance.

To try to fulfil the third MR assumption (exclusion restriction), radial MR was applied separately for each investigated outcome to detect and subsequently exclude outlying and influential genetic variants in combination with further techniques described in the next section.

### Two-sample MR, sensitivity and mediation analyses (two-step and multivariable MR)

Using the previously selected IV and the respective sample I and sample II as described above, we performed MR on the association Mapuche ancestry → GBC risk as the primary analysis. In addition to radial MR, we (1) visually inspected the funnel and scatter plots of the IV outcome (GBC) against IV exposure (proportion of Mapuche ancestry) association statistics, both adjusted for age and sex, to detect weak instrument bias; (2) calculated Cochran’s *Q* statistic using first-order inverse variance weights to detect heterogeneity, which indicates a possible violation of the instrumental variable or modelling assumptions, of which pleiotropy is a likely major cause; and (3) used the p-value for non-zero MR-Egger intercept to assess horizontal pleiotropy. Our primary objective was to identify statistical causal effects, which requires weaker MR assumptions than estimation of their magnitude, and we tested causality based on random-effect inverse variance-weighted (IVW) p-values. As a secondary objective we estimated the causal effect sizes and assessed their robustness by comparing IVW, weighted median and MR-Egger regression parameter estimates (ORs for GBC and gallstone disease, and beta values for BMI per 1% increase in the proportion of Mapuche ancestry).

Population stratification is particularly relevant in MR studies of genetically admixed individuals. To deal with potential stratification, association statistics are typically adjusted for the main principal components of genetic variability. However, the individual proportion of Mapuche ancestry was the exposure of interest in this study of Chileans, who show continuous gradients of ancestry. To check the sensitivity of MR results to population stratification, we estimated the genetic principal components in sample I and sample II using the *eigenstrat* function (Patterson et al., 2006); examined the correlation between the estimated principal components and the proportion of Mapuche ancestry; adjusted the association statistics for (1) all of the first ten principal components or (2) the first ten principal components with the exception of the principal component correlated with the proportion of Mapuche ancestry; and repeated the MR analyses.

Since a causal effect of gallstones and BMI on GBC risk has recently been reported (Barahona Ponce et al., 2021), we investigated the mediating effects of gallstone disease and BMI as a surrogate marker for obesity, as potential mechanisms explaining the relationship between Mapuche ancestry and GBC risk. The unavailability of large prospective Chilean datasets with complete information on GBC, gallstone, BMI and individual genotypes precluded the implementation of formal mediation analyses, and we decided first to apply two-step MR to assess mediation (Zheng et al., 2017). In the first MR step, IV for the proportion of Mapuche ancestry were used to assess the causal effect of the Mapuche ancestry proportion on the potential mediator (gallstone disease or BMI) by MR as described above. The second MR step was based on published findings, also based on genetically admixed Chileans, on the causal effect of the mediators on GBC risk. Evidence of association in the two MR steps (Mapuche ancestry → gallstone disease and gallstone disease → GBC) would imply some degree of mediation between Mapuche ancestry and GBC risk on the part of the intermediate trait (gallstones). We also performed MVMR to simultaneously assess the causal effects of Mapuche ancestry and gallstones on GBC risk. MVMR allows the estimation of mediation effects when considering potentially correlated risk factors utilizing genetic instruments within the MR framework. We used the R package ‘MVMR’, following the workflow suggested by the software developers (Sanderson et al., 2021). We calculated F statistics to monitor instrument strength, Q statistics to assess instrument validity including horizontal pleiotropy, and finally estimated the direct effects of the considered exposures (Mapuche ancestry and gallstone disease) simultaneously on the outcome of interest (GBC risk). The IV used included the previously selected IV for Mapuche ancestry along with five genetic variants (one palindromic variant was removed) robustly associated with gallstone disease in Chileans (Joshi et al., 2016). The sample I and sample II used to assess the association between IV and Mapuche ancestry, and between IV and gallstone disease, did not overlap. They consisted of 1,703 Chileans (1,861 minus those used as controls in sample II) and 351 gallstone patients and matched controls, respectively. The sample used to assess the direct effect between IV and gallstone disease on GBC risk was the same as the sample II utilized for the MR analysis of Mapuche ancestry → GBC risk (412 Chilean GBC cases and 412 matched controls).

MR analyses were conducted using the R version of MR-Base, which provides convenient tools for the harmonization of the association statistics, including standardization of the effect alleles and removal of problematic palindromic genetic variants (Hemani et al., 2018). The R package ‘RadialMR’ was used for radial MR (Bowden et al., 2018). For general data processing we used Plink version 1.9 (Purcell et al., 2007), and the R software environment for statistical computing and graphics (version 3.6.2). The source code to reproduce all the results described is provided as supplementary material, and the necessary input files are available at www.biometrie.uni-heidelberg.de/StatisticalGenetics/Software_and_Data.

## Data Availability

The source code to reproduce all the results described is provided as supplementary material, and the necessary input files are available at www.biometrie.uni-heidelberg.de/StatisticalGenetics/Software_and_Data.

www.biometrie.uni-heidelberg.de/StatisticalGenetics/Software_and_Data

## Funding

This study was supported by the European Union’s Horizon 2020 research and innovation program (grant 825741); the Deutsche Forschungsgemeinschaft (DFG; grant LO 1928/11-1, project number 424112940); and the Biobank of the University of Chile (BTUCH). For the publication fee we acknowledge financial support from the DFG within the funding programme „Open Access Publikationskosten“ and from Heidelberg University. Part of the research was conducted using the Hispanic Community Health Study/Study of Latinos (HCHS/SOL, dbGaP accession number phs000810.v1.p1). The HCHS/SOL study was supported by the National Heart, Lung, and Blood Institute (NHLBI) at the University of North Carolina (N01-HC65233), University of Miami (N01-HC65234), Albert Einstein College of Medicine (N01-HC65235), Northwestern University (N01-HC65236) and San Diego State University (N01-HC65237). The funders had no role in the design and conduct of the study; the collection, management, analysis, and interpretation of the data; the preparation, review, or approval of the manuscript; or the decision to submit the manuscript for publication.

## Conflict of Interest Statement

The authors declare that they have no conflicts of interest.

## Acknowledgements

The authors gratefully acknowledge the data storage service SDS@hd supported by the Ministry of Science, Research, and the Arts Baden-Württemberg (MWK) and the German Research Foundation (DFG) through grants INST 35/1314-1 FUGG and INST 35/1503-1 FUGG, and the access to computing facilities provided by Steven J. Lubbe at Northwestern University - Feinberg School of Medicine.

## Notes

### Competing Interest Statement

The authors have declared no competing interest.

### Funding Statement

This study was supported by the European Union’s Horizon 2020 research and innovation program (grant 825741, JLB, https://ec.europa.eu/info/research-and-innovation/funding/funding-opportunities/funding-programmes-and-open-calls/horizon-2020_en) and the Deutsche Forschungsgemeinschaft (DFG grant LO 1928/11-1, project number 424112940, JLB, https://www.dfg.de/en/research_funding/index.html). The funders had no role in study design, data collection and analysis, decision to publish, or preparation of the manuscript.

### Author Declarations

The study protocol conformed to the ethical guidelines of the 1975 Declaration of Helsinki and was approved by the ethics committees of Servicio de Salud Metropolitano Oriente, Santiago de Chile (#06.10.2015, #08.03.2016 and #12.11.2019), Servicio de Salud Metropolitano Sur Oriente, Santiago de Chile (#15.10.2015 and #05.04.2018), Servicio de Salud Metropolitano Central, Santiago de Chile (#1188-2015), Servicio de Salud Coquimbo, Coquimbo, Chile (#01.04.2016), Servicio de Salud Maule, Talca, Chile (#05.11.2015), Universidad Cat#x00F3;lica del Maule, Talca, Chile (#102-2020), Servicio de Salud Concepción, Concepción, Chile (ID: 16-11-97 and ID:19-12-111), Servicio de Salud Araucanía Sur, Temuco, Chile (#10.02.2020), Servicio de Salud Valdivia, Valdivia, Chile (ID:438), Centro de Bioética, Universidad del Desarrollo, Cl#x00ED;nica Alemana de Santiago, Santiago de Chile (#2018-97, ID 678) and Unidad de Investigación Hospital San Juan de Dios, Santiago de Chile (#6182), the Medical Faculties of Universidad de Chile (approval #123-2012 and #11.10.2012) and Pontificia Universidad Cat#x00F3;lica de Chile (#11-159), and Universidad de Tarapacá and University College London as described in (Ruiz-Linares et al., 2014). All participants provided written informed consent before enrolment.

